# Systems-level immunomonitoring from acute to recovery phase of severe COVID-19

**DOI:** 10.1101/2020.06.03.20121582

**Authors:** Lucie Rodriguez, Pirkka Pekkarinen, Tadepally Lakshmikanth, Ziyang Tan, Camila Rosat Consiglio, Christian Pou, Yang Chen, Constantin Habimana Mugabo, Anh Nguyen Quoc, Kirsten Nowlan, Tomas Strandin, Lev Levanov, Jaromir Mikes, Jun Wang, Anu Kantele, Jussi Hepojoki, Olli Vapalahti, Santtu Heinonen, Eliisa Kekäläinen, Petter Brodin

## Abstract

The immune response to SARS-CoV2 is under intense investigation, but not fully understood att this moment. Severe disease is characterized by vigorous inflammatory responses in the lung, often with a sudden onset after 5–7 days of stable disease. Efforts to modulate this hyperinflammation and the associated acute respiratory distress syndrome, rely on the unraveling of the immune cell interactions and cytokines that drive such responses. Systems-level analyses are required to simultaneously capture all immune cell populations and the many protein mediators by which cells communicate. Since every patient analyzed will be captured at different stages of his or her infection, longitudinal monitoring of the immune response is critical. Here we report on a systems-level blood immunomonitoring study of 39 adult patients, hospitalized with severe COVID-19 and followed with up to 14 blood samples from acute to recovery phases of the disease. We describe an IFNγ – Eosinophil axis activated prior to lung hyperinflammation and changes in cell-cell coregulation during different stages of the disease. We also map an immune trajectory during recovery that is shared among patients with severe COVID-19.

**HIGHLIGHTS:** Systems-level immunomonitoring from acute to recovery in severe COVID-19

An IFNγ - Eosinophil axis involved in lung hyperinflammation

Cell-cell coregulation differ during four disease stages

Basophils and hyperinflammation modulate humoral responses

A shared trajectory of immunological recovery in severe COVID-19

## INTRODUCTION

Since its emergence in December 2019 the Severe Acute Respiratory Syndrome-Corona Virus 2 (SARS-CoV2) causing the disease COVID-19 has infected millions of individuals and caused hundreds of thousands of deaths worldwide. The betacoronavirus has high degree of sequence homology with previous SARS-CoV and MERS coronaviruses and bind to the angiotensin-converting enzyme 2, ACE2-receptor to enter cells in the respiratory and intestinal epitelium (Lu et al., 2020).

Cells recognize the presence of the virus and its RNA via cytosolic and endosomal Pathogen recognition receptors, PRR and elicit antiviral response programs (Medzhitov and Janeway, 2002). The two main components of such antiviral programs involve the production of type I and III Interferons that induce downstream transcription of hundreds of interferon-stimulated genes that interfere with viral replication in the cell (Lazear et al., 2019). The second element of the antiviral response program is the secretion of chemokines that recruit specialized cells of the immune system to clear the virus. SARS-CoV-2, like other viruses, has evolved countermeasures to these defenses and in particular, the virus efficiently interferes with IFN-signaling and the induction of ISGs in SARS-CoV-2 infected cells (Blanco-Melo et al., n.d.). In contrast, pro-inflammatory cytokine and chemokine responses are induced normally and this imbalance between antiviral and pro-inflammatory responses is a key feature of COVID-19 (Vabret et al., 2020).

Another observation during the COVID-19 pandemic is the different disease courses among different individuals infected by SARS-CoV2 virus. Most individuals present with a very mild disease, often asymptomatic, and a few develop a life-threatening disease requiring intensive care. The strongest determinant of disease severity is age, with children presenting almost exclusively with mild disease (Brodin, 2020), while the elderly, over 70 years of age are much more likely to develop severe COVID-19. Males and females are infected at similar rates, but males are much more likely to develop severe disease and requiring intensive care (Jin et al., 2020). Obesity, smoking and hypertension are other risk factors for severe COVID-19 (Huang et al., 2020). On the other hand, COVID-19 contrast with other respiratory viral infections in that pregnant women do not seem to be more likely to develop severe disease and this is also true for patients with various forms of immunodeficiency. One likely reason for these observations is that severe disease is associated with exuberant immune responses and a skewed immune regulation against pro-inflammatory responses in pregnancy and T-cell deficiencies in transplan patients make such hyperinflammatory responses less likely. To treat hyperinflammation in severe COVID-19 we need to better understand the cells involved, their interactions and protein mediators used to orchestrate their response. To this end, we have perfomed systems-level analyses of the immune system in blood from 39 patients, from acute to recovery phase of severe COVID-19, with up to 14 blood samples collected from a given patient. These analyses reveal a sequence of responses involving many immune cell populations at different stages of the disease. A transient response involving IFNγ upregulating CD62L on Eosinophils prior to lung hyperinflammation, examples of coregulated cell populations, and immune correlates of productive antibody responses to SARS-CoV2 as well as an integrated immune trajectory shared across patients recovering from severe COVID-19.

## RESULTS

### Longitudinal profiling of patients with COVID-19

Given the enormous diversity among immune systems in humans, longitudinal monitoring of patients is required to appreciate immunological changes occurring during a disease process. Also, systems-level analyses methods such as Mass cytometry (Brodin, 2018) enable all immune cell populations to be distinguished and analyzed in a given blood sample, allowing for coordinated changes across cell populations to be revealed. We have combined these cellular measurements with analyses of 180 unique plasma proteins using Olink analyses (Lundberg et al., 2011) (**Fig. 1a**). In order to understand systems-level immune responses during moderate to severe COVID-19 we monitor longitudinal samples from 39 patients, some treated in the intensive care unit (ICU) and some treated in regular hospital wards with oxygen support but no mechanical ventilation (**Fig. 1b**). Patients did not receive immunomodulatory therapies in this cohort and the immunological changes reflect the natural course of the infection and all patients survived the infection.

**Figure 1.**
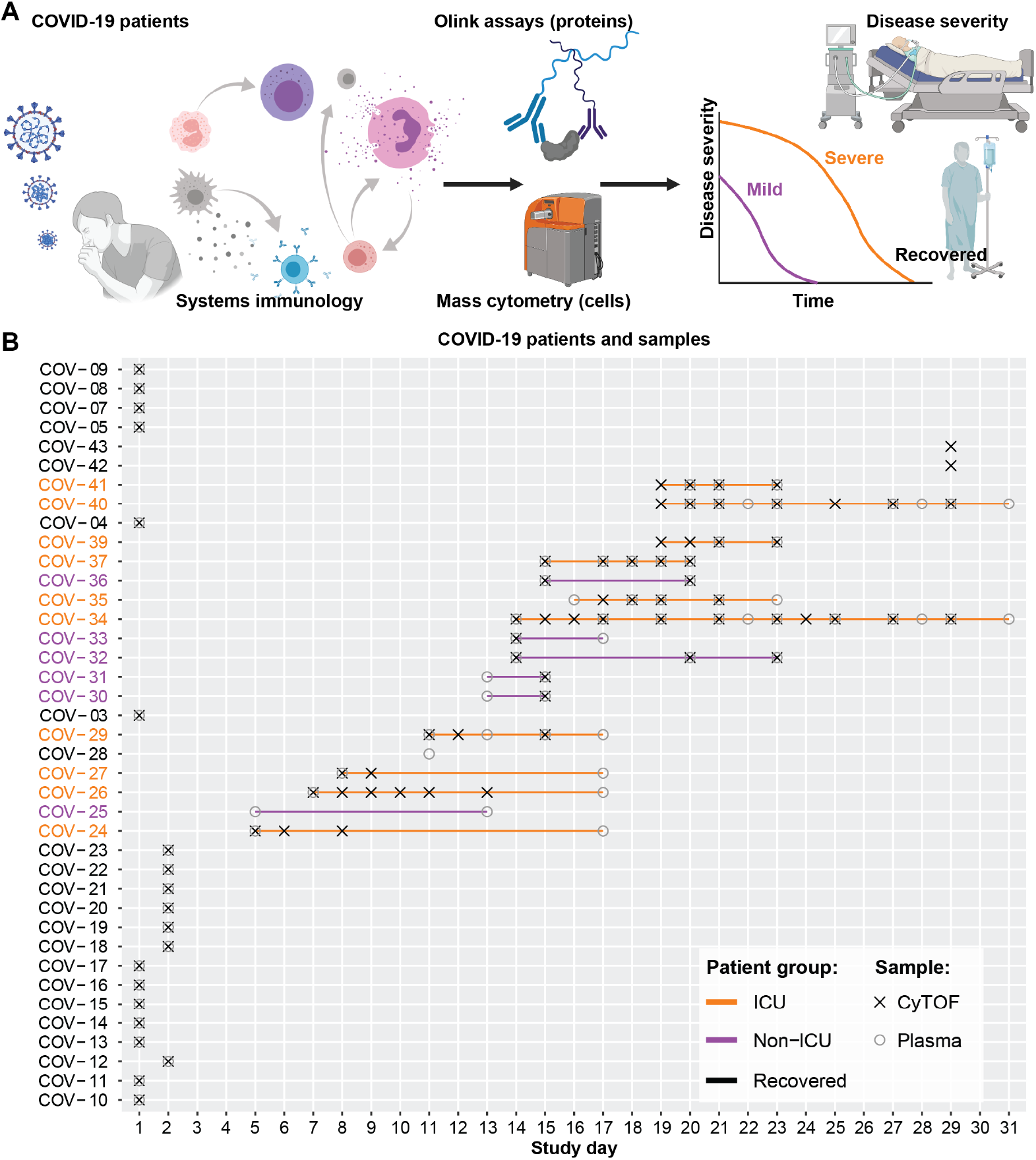
Longitudinal profiling of the immune system in moderate and severe COVID-19. (**A**) 180 unique plasma proteins were quantified using Olink assays (n = 77 plasma samples) and whole blood immune cells analyzed by Mass cytometry (n = 80 whole blood samples). (**B**) Monitoring and longitudinal sampling of blood cells (X) and plasma (o) from 39 patients at the Helsinki University Hospital.

### The characteristics of acute and recovery phases of COVID-19

Clinical measurements were taken from acute and recovered patients including body temperature, white blood cell counts and lymphocyte counts. Milder cases of COVID-19 showed lower body temperatures as well faster normalization of body temperatures as compared to severe cases who fluctuated more over time (**Fig. 2A**). The white blood cell (WBC) counts gave a possible correlate to the stage of the infection. High WBC counts are often a reflection of acute inflammation and immune responses and in severe patients we observed more fluctuating levels of WBC over time (**Fig. 2B**). Also, there were no signs of secondary bacterial infection in the patients in this cohort. Lymphopenia is one of the common features of COVID-19 and the degree of lymphopenia predict disease severity (Huang et al., 2020). Lymphocyte counts were measured and found that milder cases recuperated their lymphopenia faster than severe cases (**Fig. 2C**). This is in line with other previous reports (Lagunas-Rangel, 2020).

**Figure 2.**
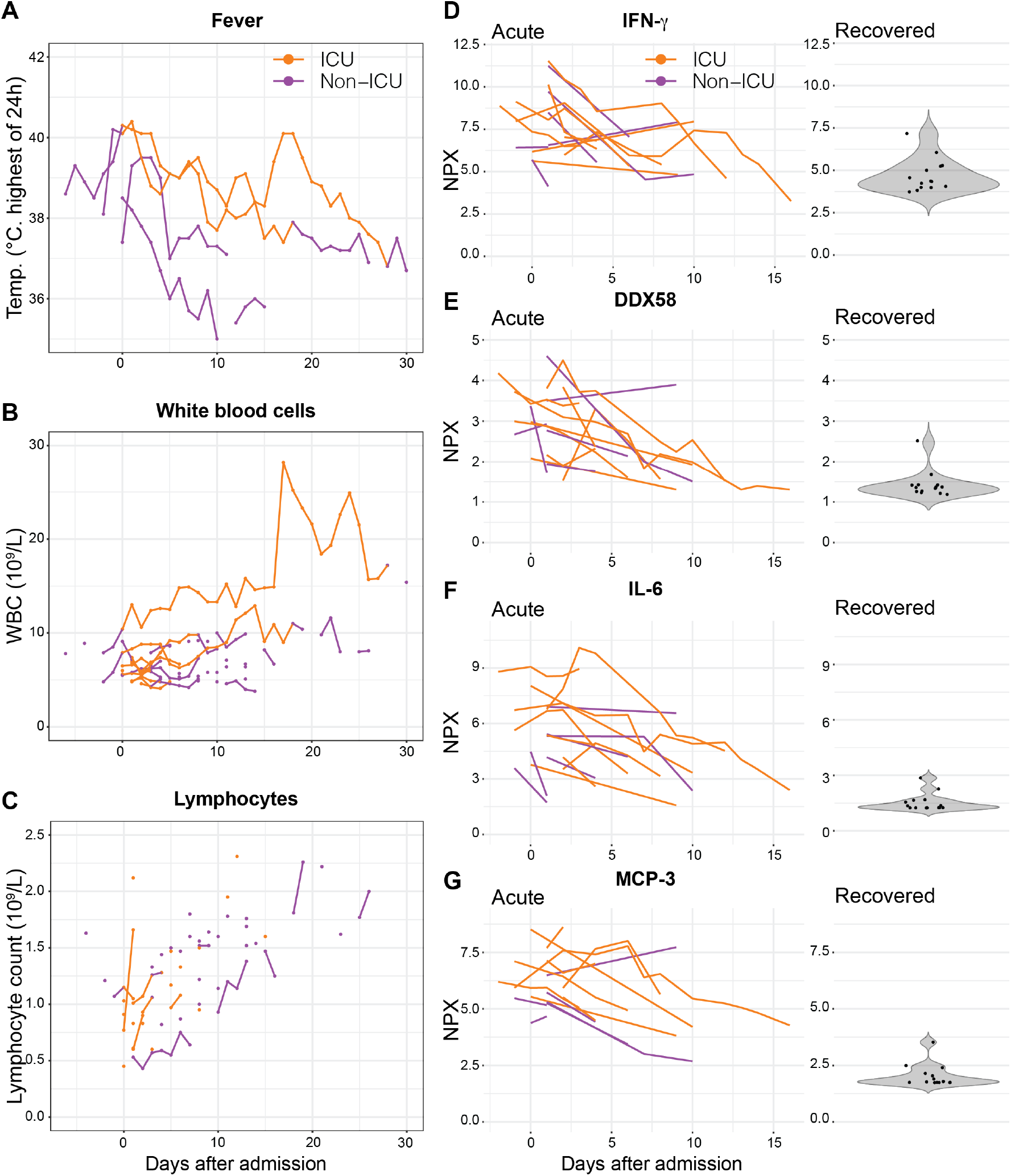
The natural course of severe COVID-19 from admission to clinical recovery. (**A**) Body temperature measurements from representative patients over the course of 30 days from admission to the hospital in ICU and non-ICU patients. (**B-C**) White blood cell counts, and Lymphocyte counts during acute and recovery phase in COVID-19 patients. (**D-G**) Plasma levels of the indicated proteins using Olink assays in longitudinal samples from acute patients (left) and single measurements from recovered patients (right). NPX: Normalized Protein expression.

Plasma protein levels were observed and compared among acute and recovered phases and map immune dynamics of severe COVID-19 (**Fig. 2D-G**). Pro-inflammatory cytokines like IL-6 and IFNγ predict disease severity. A decreasing trend is observed in IFNγ and IL-6 from early admission to the hospital through recovery during the weeks of the study (**Fig. 2D and 2F, respectively**). Similarly, DDX58, the Innate Immune Response Receptor also called RIG-I, and the monocyte chemoattractant protein MCP-3, are elevated during acute disease and decrease during recovery (**Fig. 2E and 2G, respectively**).

### The immune cell changes from acute to recovery phase of COVID-19

A defining feature of acute immune responses during COVID-19 is dramatic changes in immune cell composition that can be informative of likely driving factors and triggers. To understand severe COVID-19 better we plot relative proportions of 57 immune cell populations over time in the 39 patients (**Fig. 3**). We confirm the overrepresentation of Neutrophils over Lymphocytes during acute infection, that is slowly reversed during the recovery phase (**Fig. 3**). This is in line with reports suggesting the Neutrophil-to-Lymphocyte ratio (NLR) and degree of lymphopenia are predictive of disease severeity in COVID-19 (Lagunas-Rangel, 2020). The plasmablast response is clear and occurs during the first week after admission in these patients (**Fig. 3**). The recovery of T-cells after the initial lymphopenia occurs over the following 2–3 weeks and is dominated by CD127 expressing effector and central memory CD4^+^ T-cells, as well as CD57-expressing and differentiated memory CD8^+^ T-cells (**Fig. 3**). Also, all Dendritic cell (DC) subsets increased from acute to recovery phases, CD1c^+^ DCs, CD11c^+^ DCs and plasmacytoid DCs (pDC) (**Fig. 3**). Also, despite a clear reduction in relative abundance of neutrophils over time, the other granulocyte subsets, Basophils and Eosinophils increased clearly from acute to recovery phases (**Fig. 3**), and both of these were among the most dynamic cell populations during severe disease, suggestive of important contributions to the antiviral defense and immunopathology.

**Figure 3.**
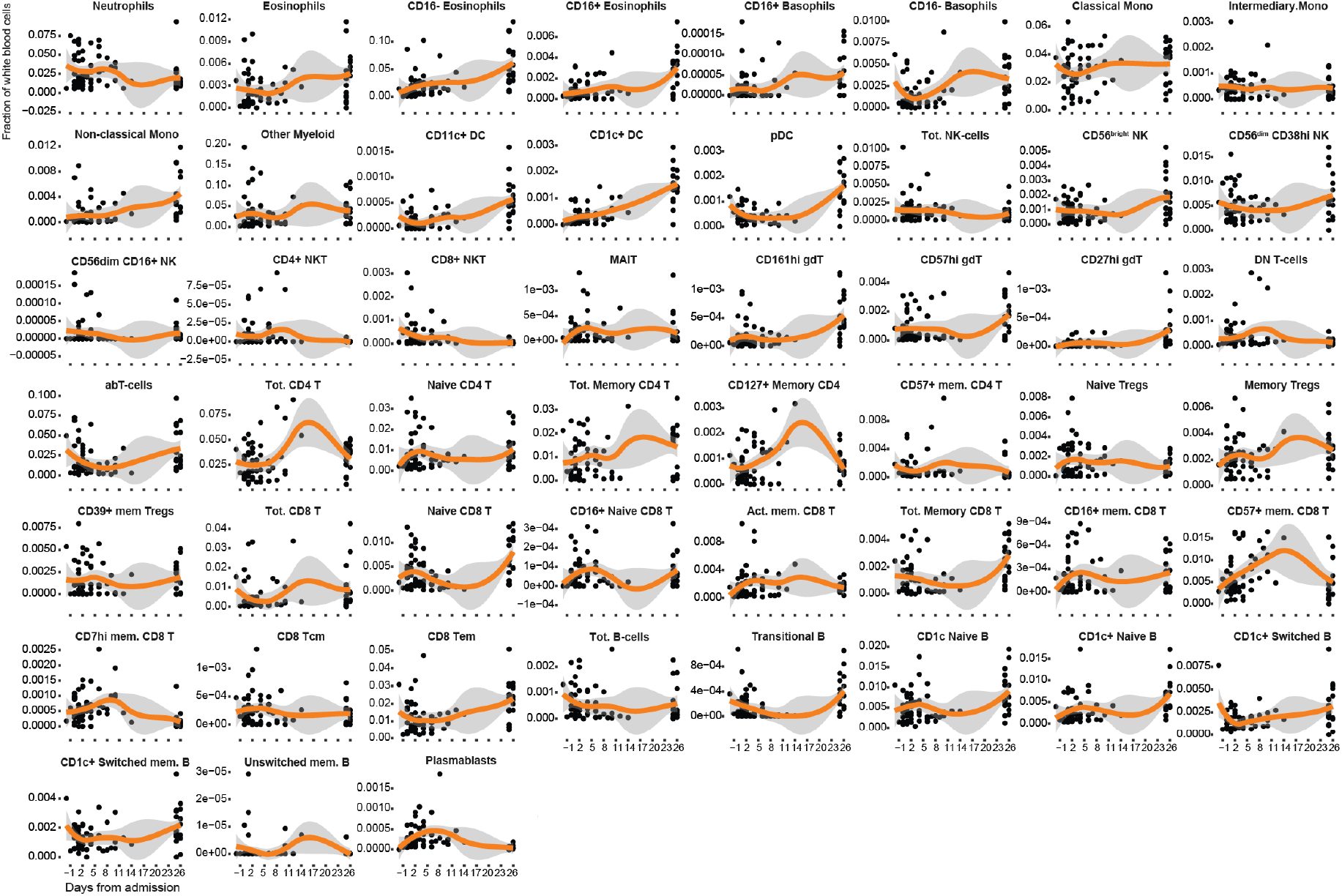
Immune cell proportions in COVID-19. Proportion of 57 white blood cell populations determined by mass cytometry. Loess smoothing in orange.

### Eosinophil activation and homing during acute COVID-19

Given the changes in eosinophil abundance above, we decided to study Eosinophils more carefully. There are reports of strong GM-CSF responses in lungs of COVID-19 patients (Zhou et al., 2020) and GM-CSF is known to stimulate Eosinophils, particularly in interstitial pneumonia and allergic inflammation (Taniguchi et al., 2000). Taking advantage of the detailed longitudinal sample series, we used Partition-based graph abstraction, PAGA (Wolf et al., 2019), to reconstruct single-cell phenotypic changes of blood eosinophils during acute COVID-19 (**Fig. 4**). Leiden clustering found 12 Eosinophils subsets and the main groups are annotated by defining features (**Fig. 4A)**. By splitting cells obtained from the different longitudinal samples, time-associated changes in eosinophil phenotypes are revealed with a transient expansion of CD62L^+^ Eosinphils from day 2 to day 6 after admission (**Fig. 4B)**. CD62L-upregulation on Eosinophils have been reported to be induced by IFNγ(Momose et al., 1999), one of the most elevated cytokines in severe COVID-19, and the IFNγ levels show a slight increase right around the same time as the expansion of CD62L+ Eosinophils (**Fig. 4C)**. This phenotype of Eosinophils is reminiscent of lung resident Eosinophils, rather than induced inflammatory eosinophils in circulation and such ung-homing cells have previously been reported as important homeostatic regulators of inflammatory responses in the lung (Mesnil et al., 2016) (**Fig. 4D**). It is tempting to think that this transient expansion of CD62L+ Eosinophils just prior to the time development of severe lung hyperinflamation around 1 week after admission is related to this immunopathology of the lungs in COVID-19 patients. To this end further investigation into this Eosinophil – IFNγ axis is required and might suggest novel therapies targeting this response to mitigate ARDS and lung inflammation.

**Figure 4.**
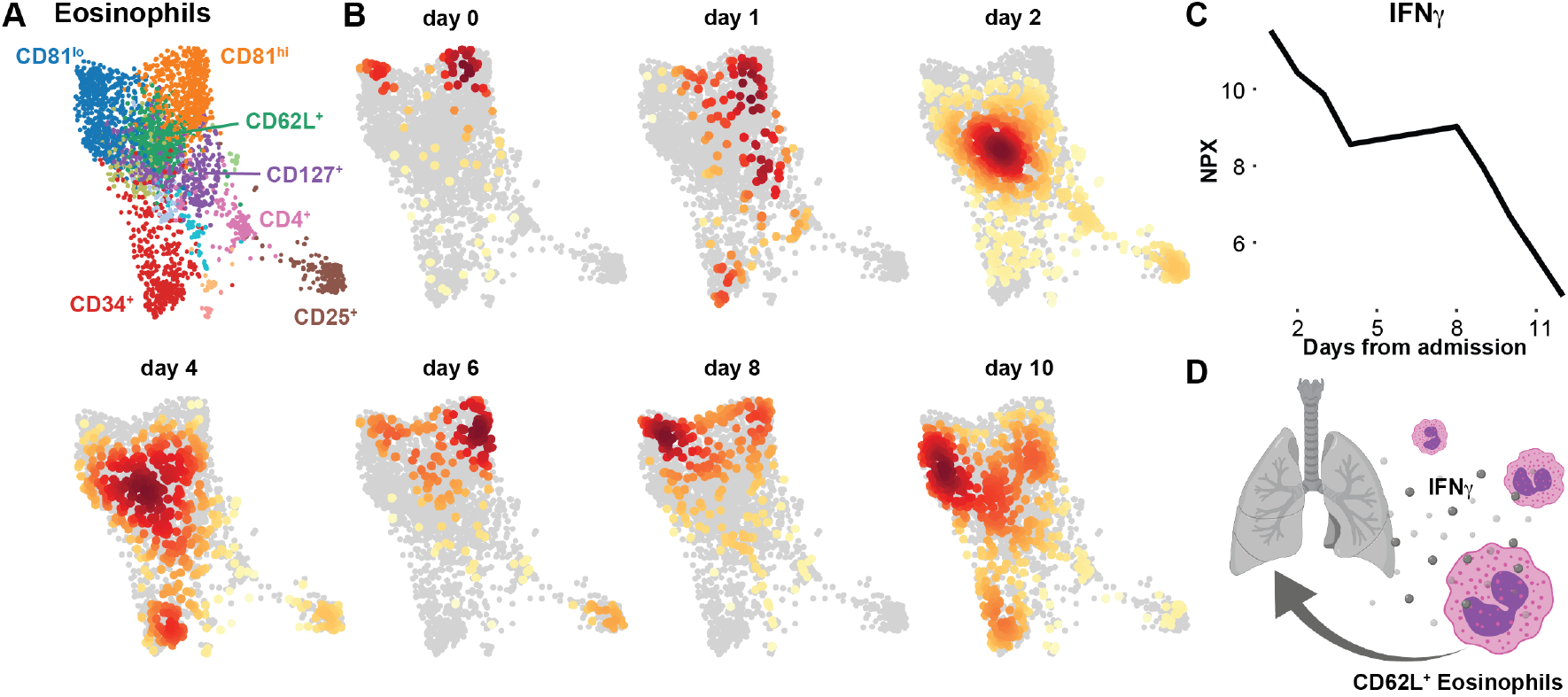
Eosinophil changes from admission to recovery. 2D representation generated by Partition-based graph abstraction (PAGA) of Eosinophils from patient COV-40 at 7 different timepoints from admission to recovery. (**A**) Louvain clusters are colored and annotated by key protein characteristic and (**B**) cell distributions at each individual timepoint indicate changes in immune cell states and composition over time. (**C**) Plasma IFNγ levels as measured by Olink assay in plasma samples from the same patient COV-40. (**D**) IFNγ mediated upregulation of CD62L contributes to lung inflammation hyperinflammation.

### Adaptive immune cell dynamics during severe COVID-19

Adaptive responses to SARS-CoV2 are seen in most individuals, with one study reporting CD4^+^ T-cell responses, and CD8^+^ T-cell response in nearly all patients (Grifoni et al., 2020). Similarly, the majority of symptomatic patients seroconvert within a few days and most developed high-titer antibody responses (Sun et al., 2020), yet one study has reported that a significant proportion of patients with COVID-19 do not develop neutralizing antibody responses (Robbiani et al., 2020). To investigate the dynamics of adaptive immune cell responses in our cohort we used the same PAGA approach as described above. We find a clear Plasmablasts response early after admission (**Fig. 5A**). The CD4^+^ T-cell response were initially dominated by effector and central memory responses, followed by an increase in Tregs approximately four days after admission (**Fig. 5B**). The CD4^+^ T-cells were split into two effector cell populations based CD4-expression level, possible reflecting an activation-induced downregulation in a subset of CD4^+^ T-cells (**Fig. 5B**)(Grishkan et al., 2013). The CD8^+^ T-cell responses are dominated by activated cells expressing high CD38 and also a subset of effector cells upregulating the CD147 receptor from about one week onwards (**Fig. 5C**). Gamma-delta TCR T-cells, (γδT-cells) and CD8^+^ T-cells progressively upregulated the marker of terminal maturation CD57 from about 1 week onwards (**Fig. 5C-D**). These results are largely in agreement with other recent reports (Mathew et al., 2020) and highlight the strong innate and adaptive immune activation during acute COVID-19.

**Figure 5.**
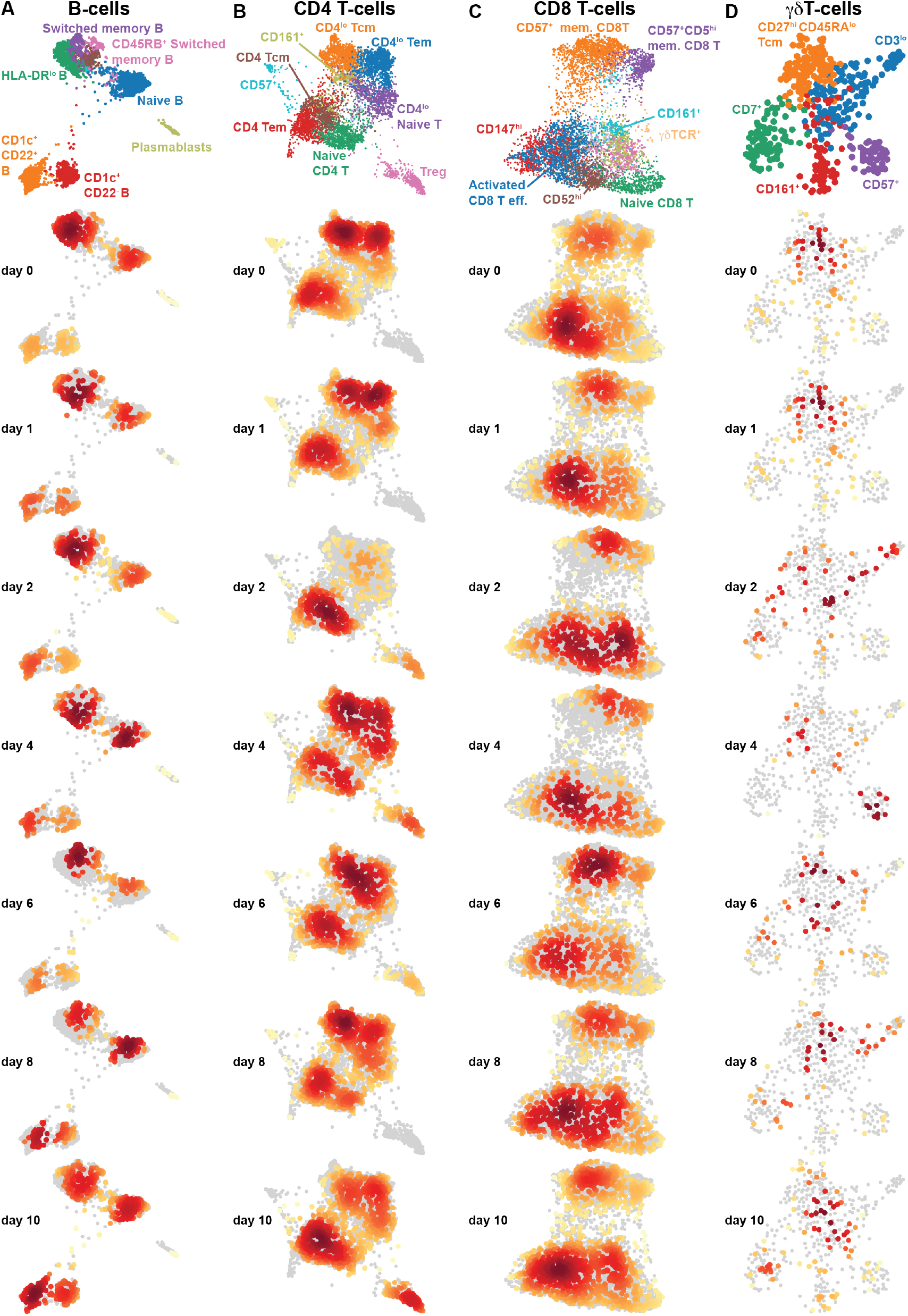
Adaptive immune cell changes from admission to recovery. (**A-D**) 2D representation generated by Partition-based graph abstraction (PAGA) B-cells, CD4 T-cells, CD8 T-cells and γδT-cells from patient COV-40 at 7 different timepoints from admission to recovery. Louvain clusters are colored (top) and annotated by key protein characteristic and cell distributions at each individual time point indicate changes in immune cell states and composition over time.

### Cell-cell regulation varies over time during severe COVID-19

Immune responses are always concerted efforts made by multiple, specialized cell populations communicating via direct interactions and secreted cytokines and other mediators. By studying such cell-cell relationships a better understanding of the systems-level response can be obtained. We generated cell-cell correlation matrices using longitudinal cell population frequencies and binned the samples into four phases from acute disease to recovery phase (**Fig. 6A**). We find that the first phase (day 0–4) is dominated by an inverse correlation between neutrophils and a number of myeloid and lymphoid cell types, as reflected in the elevated NLR, associated with severe disease (Lagunas-Rangel, 2020)(**Fig. 6A**). The following phase (day 6–8) is characterized by a strong coordinated Plasmablast, B-cell and abT-cell module, and this is inversely correlated with a strong Treg and CD11c^+^ DC module (**Fig. 6A**). From day 9 onwards a change is apparent with a shift towards a coregulated module involving Eosinophils, pDCs, CD11c^+^ DCs, with CD8^+^ T-cells. This module is largely maintained in the recovered patients, possibly reflecting a more normal cell-cell relationship (**Fig. 6A**).

**Figure 6.**
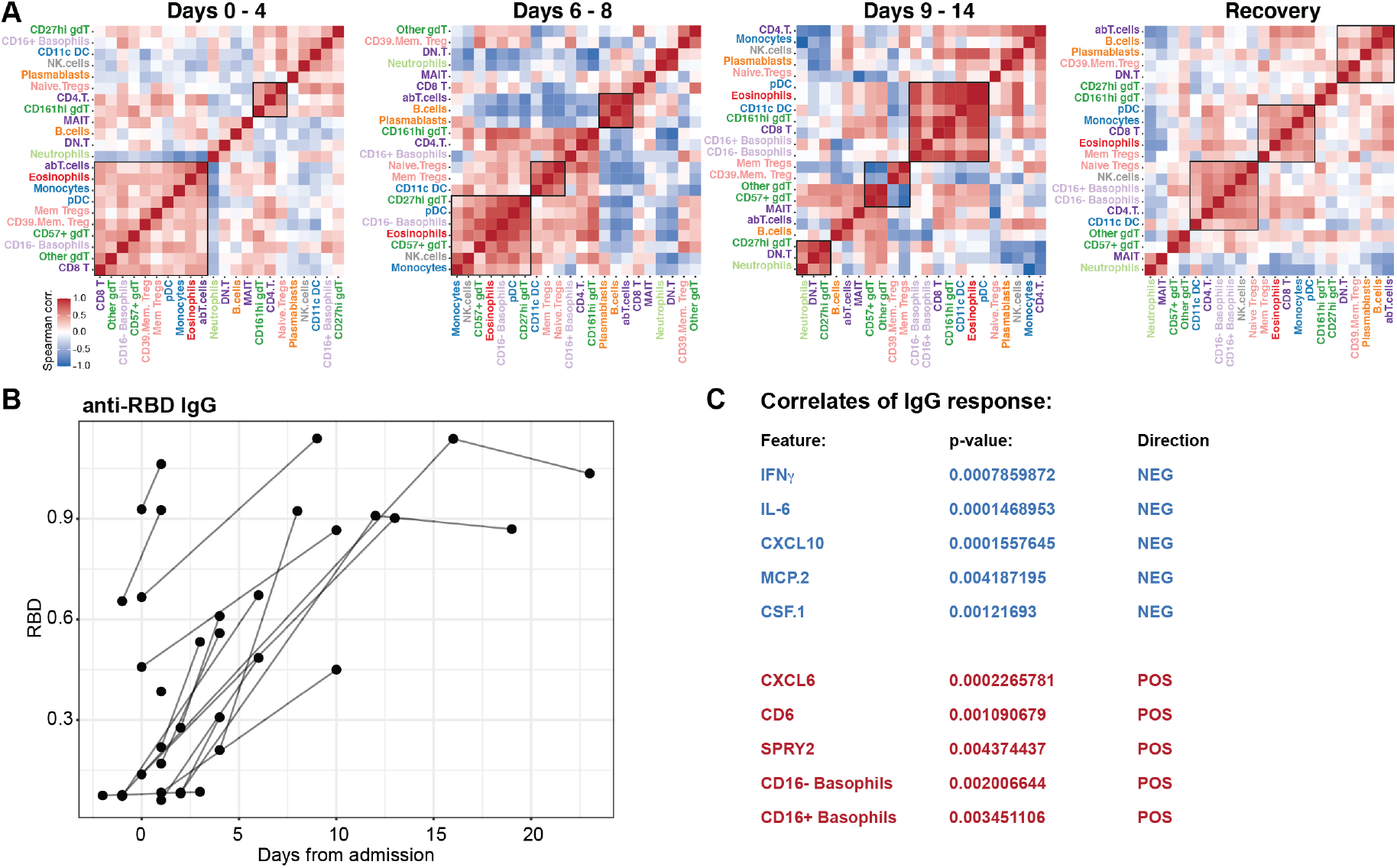
Cell-cell communications network during different phases from acute to recovery of COVID-19. (**A**) Spearman correlation matrices from samples collected at the indicated time intervals and ordered by top correlations. Co-regulated cell populations highlighted by boxes. (**B**) Serum IgG antibodies against SARS-CoV2 Spike protein Receptor Binding Domain, RBD showed against days after admission. (**C**) Mixed effect modelling, MEM of plasma protein levels and immune cell population frequencies against anti-RBD IgG titers. Five most positively and negatively associated features in MEM correlated with antibody responses when Days from admission is taking into account as a fixed effect.

A prototype of a coordinated immune response to viruses is the appearance of virus specific IgG antibodies because such responses elicited by B-cells require help from CD4^+^ T-cells. Here we investigated the seroconversion in this cohort and found strong induction of IgG antibodies to the SARS-CoV2 Spike protein (Receptor Binding Domain, RBD) in the majority of patients (**Fig. 6B**). This is in line with similar analyses in other COVID-19 patients (Sun et al., 2020)(Amanat et al., 2020). We were unable to test the neutralizing capacity of these antibodies at this time, but another recent report has shown that a significant proportion of patients mount antibodies that lack such neutralizing capacity (Robbiani et al., 2020). To understand the immunological correlates of IgG responses to SARS-CoV2, we devised a mixed effect model, using both plasma protein levels and cell frequencies as predictors, taking days after admission into account as a fixed effect (Chung et al., 2013). We found several features significantly associated with IgG-responses, and in particular strong proinflammatory cytokines IFNγ, IL-6, and chemokines CXCL10 and MCP-2 (CCL8) are negatively associated with anti-CoV2 IgG responses (**Fig. 6C**). In contrast the Neutrophil-recruiting chemokine CXCL6 are positively associated with anti-CoV2 IgG responses and so was the fraction of circulating Basophils (**Fig. 6C**). It is known that basophils are able to bind antigens on their surface and potentiate humoral immune responses (Denzel et al., 2008) and since basophils are depleted during acute and severe COVID-19 (**Fig. 3**), our data collectively suggest that the degree of basophil depletion might influence the efficacy of IgG-responses to SARS-CoV2. It is believed that basophil mediated enhancement of B-cell responses occurs through the production of either IL-4 or IL-6, but levels of the latter were found to inversely associated with antibody responses (**Fig. 6C**) so it more likely that another mechanism is responsible for the basophil enhancement of IgG responses in the case of COVID-19. Collectively these results indicate a coordinate adaptive immune response to SARS-CoV2, enhanced by basophils and possibly suppressed by hyperinflammatory cytokine responses with high IL-6 levels during acute COVID-19.

### A shared, integrated trajectory of recovery across patients

Since none of the patients in this cohort were treated with immunomodulatory agents, and have recovered with supportive care alone, we reasoned that a deeper investigation into the immunological changes during recovery from severe COVID-19 would be informative about the underlying immune processes involved. Given the strong interactions among immune cells and proteins in the immune system we applied an integrative analysis method to search for a multiomic trajectory of immune recovery. We used Multiomics Factor Analysis, MOFA (Argelaguet et al., 2018). This method allowed us to search for latent factors that best explain the variance across data types and use these to discern any possible relationship with the process of recovery from disease.

We found ten latent factors that explained the variance in the combined dataset (**Fig. 7A**), and out of those, latent factor 2 was associated with the transition from acute to recovery phases of disease (**Fig. 7B**). There were no clear differences among intensive care unit (ICU) or non-ICU ward patients and the levels of latent factor 2 were highest in the samples taken from recovered patients (**Fig. 7B**). To understand the biology of immune recovery we assessed the underlying features contributing to latent factor 2. The plasma proteins changing the most all decreased during recovery and most prominent were IL-6, MCP3, KRT19 (Keratin19), CXCL10, AREG and IFNγ (**Fig. 7C**). Conversely the cells that changed the most during recovery were classical and non-classical monocytes, CD56^dim^ NK cells, Eosinophils and γδT-cells, all increasing in relative proportions during recovery (**Fig. 7D**). This shared, integrated trajectory reveal markers most indicative of recovery in patients with severe COVID-19 and if verified in independent sets of patients, these features could be valuable biomarkers to monitor during disease progression in order to detect a switch from acute to recovery phases in severe COVID-19.

**Figure 7.**
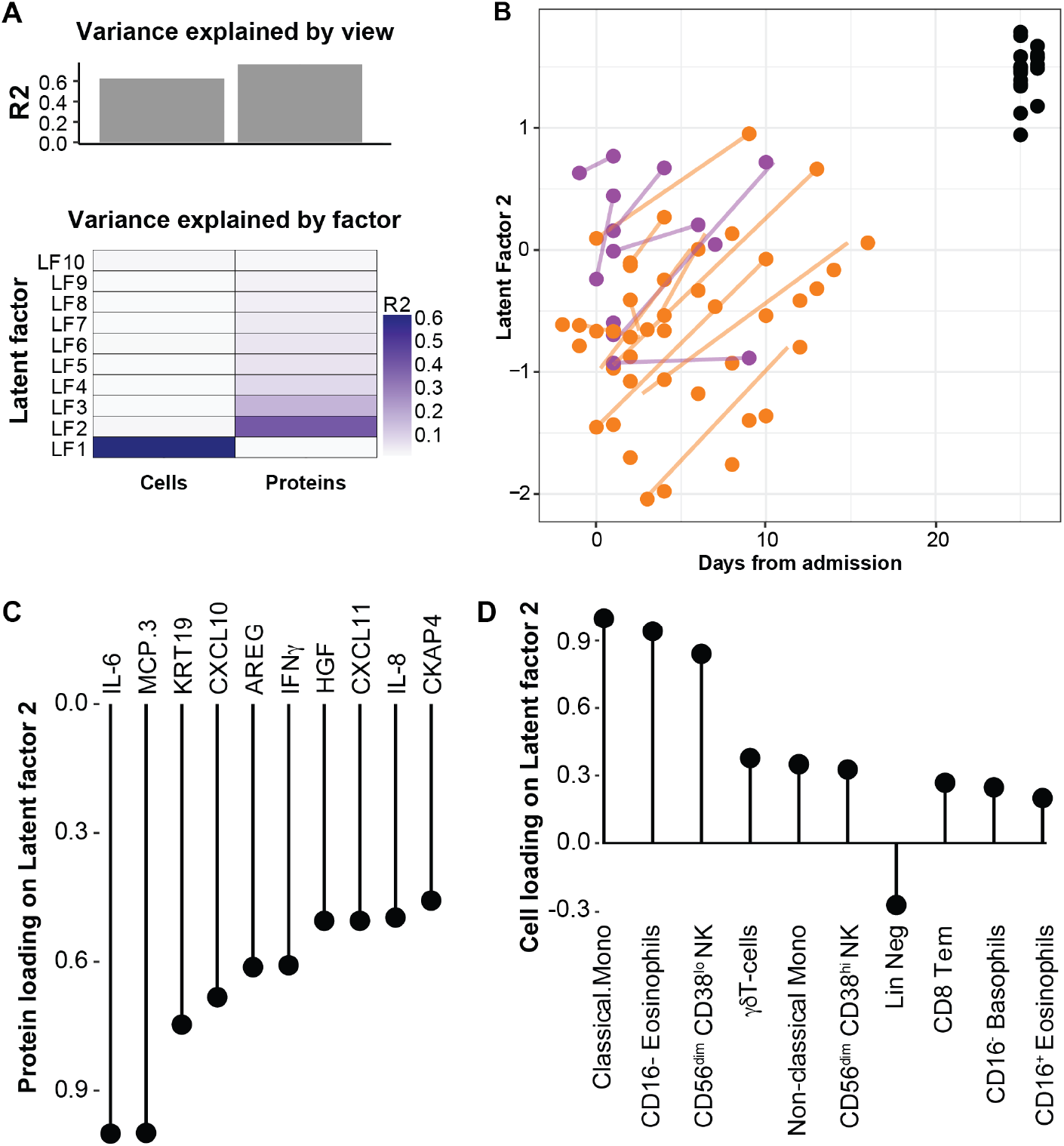
A multiomics immune signature from acute COVID-19 to recovery. Multiomics Factor Analysis, MOFA is used to integrate 148 plasma protein levels, and 63 immune cell frequencies across all 99 blood samples collected from 39 patients. (**A**) Fraction of total variance explained by type of measurement (view), and by Latent Factors, LFs 1–10. (**B**) LF2 best represent the changes from acute to recovery over time and reveal a shared trajectory for most patients. (**C**) Lollipop plot show plasma proteins explaining LF2. (**D**) Lollipop plot show cell population frequencies explaining LF2.

In this manuscript we have performed an in depth, longitudinal analysis of the immune system in patients with severe COVID-19 during acute disease and until spontaneous recovery. The natural course of this process is mapped and found to be similar among patients. We find changes in cell populations, such as CD62L-expressing Eosinophils, triggered by IFNγ and likely contributing to hyperinflammation and ARDS during acute disease. We show that basophils are depleted during acute disease but recuperate during recovery and the levels of basophils are significantly correlated with the titers of IgG antibodies to SARS-CoV2 produced by B-cells. In contrast, high levels of IL-6 and IFNγ are negatively associated with humoral responses. Finally, we uncover an immunological trajectory of disease recovery shared among patients. These results can be useful for the development of better immunomodulatory strategies to mitigate hyperinflammatory responses, optimize antiviral IgG responses and monitoring of disease progression and recovery in patients with severe COVID-19.

## DISCUSSION

A number of researchers are studying the immune response to SARS-CoV2 and we are learning that viral evasion of IFN-I/III signals and prevention of the normal induction Interferon-stimulated genes and the antiviral state (Blanco-Melo et al., n.d.). At the same time the proinflammatory response is strong. Secretion of chemokines and proinflammatory cytokines lead to influx of neutrophils and myeloid cells into the lung with strong local inflammatory responses and immunopathology (Vabret et al., 2020). Autopsy findings in patients succumbed to COVID-19 are characterized by perivascular T-cell infiltration, microangiopathy and widespread thrombosis in the lung tissue (Ackermann et al., 2020). The induction of IL-6 during severe COVID-19 have led to trials of blocking antibodies to the IL-6 receptor with mixed results. This is inspired by Cytokine Release Syndromes, CSR seen in cancer immunotherapy also often treated with IL-6 blocking agents. However, there are a number of differences between severe COVID-19 and CRS, such as lower IL-6 levels, death caused by respiratory failure and thrombosis, rather than from circulatory failure and status epilepticus as seen in CRS (Vardhana and Wolchok, 2020). In this respect the mechanisms of severe COVID-19 are incompletely understood and better understanding is required for improved immunomodulatory therapies to be devised and immunopathology and mortality limited.

Human immune systems are highly variable (Brodin and Davis, 2016), and most of this variation is explained by environmental exposures (Brodin et al., 2015), particularly early in life (Olin et al., 2018). The role of genetic variation in immune variation in general and in COVID-19 in particular is under investigation (Casanova et al., 2020). Systems-level analysis methods are useful in human immunology because they capture the many variable cell populations, proteins and transcriptional programs involved in a complex immune response. Systems-level analyses also allow for the inference of relationships among such immune system components (Lakshmikanth et al., 2020). In this study we add to the rapidly growing literature by providing a longitudinal, systems-level perspective on the immune system changes from acute to recovery phases of severe COVID-19 disease. Longitudinal analyses are important because cross-sectional analyses carry the risk of capturing snapshots of patients at different stages of the immune response and thereby misinterpret differences as qualitatively different. The longitudinal sampling presented herein is a strength of the current study. Another important aspect of this work is its use of whole blood, rather than peripheral blood mononuclear cells, allowing neutrophils, and other granulocyte populations to be included in the analysis and also reduce the technical sources of variation caused by cell preparation and freezing (Brodin et al., 2019). By using this more holistic and longitudinal approach to analyze the immune response during severe COVID-19, we find previously unappreciated roles of Eosinophils in the acute response. These cells play important roles in other respiratory infections (Flores-Torres et al., 2019) but have not been implicated much in COVID-19. The population of eosinophils that expand a few days after admission to the hospital were carachterized by CD62L-expression, a previously reported marker of lung eosinophils (Mesnil et al., 2016) and it is probable that this IFNγ-mediated upregulation of CD62L on eosinophils lead to the influx of such cells into the lung tissue and the development of ARDS and clinical deterioration often seen after about one week in many patients. The finding that basophil levels are positively associated with humoral responses to SARS-CoV2 is intriguing and in line with previous studies in other viral infections (Denzel et al., 2008). Further investigation will be required to understand the mechanisms involved but it is likely not involving the production of IL-6 by basophils given that plasma levels of this cytokine were inversely associated with IgG response titers induced. Another possible mechanism involve IL-4 production by basophils, known to potentiate B-cell responses to infection (Kawakami, 2008).

There has been a lot of concerns around antibody responses to SARS-CoV2 and although nearly all patients with severe disease do produce antibodies in rather high titers (Amanat et al., 2020; Grifoni et al., 2020), the neutralizing capability of such antibodies are variable (Robbiani et al., 2020). One hypothesis brought forward as a possible explanation to the severe disease occuring often after a week or so of stable disease is antibody-mediated enhancement (ADE)(Iwasaki and Yang, 2020; Tetro, 2020). This occurs when non-neutralizing antibodies bind a virus and via Fc-receptors bring viruses into new cell types, not expressing the receptor required forviral entry, in this case ACE2. Such reponses are well known for Dengue virus infection and could induce hyperinflammatory responses also in COVID-19. We have found that a significant proportion of CD4^+^ T-cells in some patients showed CD4 downregulation as a sign of possible cell activation, but such downregulation can also occur if T-cells are directly infected (Xiang et al., 2009). CD4^+^ T-cells do not express ACE2 (Uhlen et al., 2019), but could express Fc-receptors and therby be subject to viral infection and replication via ADE. This is speculative at this time but as more data on determinants of neutralizing antibody responses the theory of ADE as a cause of severe COVID-19 will be testable and have important implications for vaccine development (Iwasaki and Yang, 2020). The influence of basophils in modulating humoral responses to SARS-CoV2 uncovered herein should also be taken into account as basophils are depleted during acute disease and the severeity if such depletion might be an important determinant of the antibody response to the virus.

## Data Availability

All raw data and code required to reproduce all figures is available freely.

https://ki.box.com/s/sby0jesyu23a65cbgv51vpbzqjdmipr1

## ACKNOWLEDGEMENTS

The authors are grateful to private donations to Karolinska Institutet from Bure Equity AB (Stockholm, Sweden) and Jonas and Christina af Jochnick Foundation. The study was also supported by grants from Academy of Finland to E.K. (308913) and S.H. (323499) and Helsinki University Hospital (project M7100YLIT2) to P.P as well as funding from Juho Vainio Foundation to O.V and A.K. We appreciate the hard work of doctors and nurses at the Helsinki University Hospital. We thank the team at the SciLifeLab, Plasma Profiling Facility in Stockholm for generating the Olink data.

## CONTACT FOR REAGENT AND RESOURCE SHARING

All requests to

## EXPERIMENTAL MODEL AND SUBJECT DETAILS

Non-interventional, observational study.

### Ethics

The study was approved by the Ethics Committee of the Hospital District of Helsinki and Uusimaa (HUS/853/2020) and conducted in accordance with the Declaration of Helsinki. Written informed consent was obtained from all participants.

### Patients

#### Inpatients

We included symptomatic patients with positive SARS-CoV-2 PCR test admitted to Helsinki University Hospital, Helsinki, Finland. Patients were recruited within five days after hospitalization. We excluded patients who had been considered by the attending clinician not to benefit from intensive care. We recruited 17 inpatients (9 females, 8 males) aged between 40 – 77 years. The duration of hospitalization ranged from 5 to 38 days. Of these, 10 were admitted to the ICU, and remained in intensive care for 1 – 27 days. Three patients required mechanical ventilation for 3, 13 and 19 days, respectively.

#### Recovery phase patients

In addition to patients recruited during the acute phase of illness, we recruited a separate cohort of recovered patients based on positive PCR (n = 20) or high clinical suspicion (n = 2). These 22 subjects (age range 28 – 68 years; 11 females, 11 males) were included during convalescence 3–4 weeks after COVID-19 diagnosis and SARS-CoV-2 detection. These patients were identified from medical and laboratory records, contacted by phone and invited to donate a blood sample.

## METHOD DETAILS

### Immunophenotyping by Mass Cytometry

Blood samples drawn from patients with COVID-19 were mixed with a whole blood stabilizer (Brodin et al., 2019)(Cytodelics AB, Sweden) either immediately or within 1–3 hrs post blood draw and cryopreserved as per the manufacturer’s recommendations. Samples were then thawed, and cells were fixed/RBCs lysed using WASH # 1 and WASH # 2 buffers (Whole blood processing kit; Cytodelics AB, Sweden) as per the manufacturer’s recommendations. This was performed a few days prior to barcoding and staining of cells. Post fix/lysis of cells, ∼1–2×10^6^ cells/sample were plated onto a 96 well round bottom plate using standard cryoprotective solution (10% DMSO and 90% FBS) and cryopreserved at −80°C.

At the time of experimentation, cells were thawed at 37°C using RPMI medium supplemented with 10% fetal bovine serum (FBS), 1% penicillin-streptomycin and benzonase (Sigma-Aldrich, Sweden). Briefly, cells were barcoded using automated liquid handling robotic system (Agilent technologies)(Mikes et al., 2019) using the Cell-ID 20-plex Barcoding kit (Fluidigm Inc.) as per the manufacturer’s recommendations. Samples were pooled batch wise by keeping together the longitudinal samples from each patient in the same batch. Cells were then washed, FcR blocked using blocking buffer (in-house developed recipe) for 12 min at room temperature, following which cells were incubated for another 30 min at 4°C after addition of a cocktail of metal conjugated antibodies targeting the surface antigens. Cells were washed twice with CyFACS buffer (PBS with 0.1% BSA, 0.05% sodium azide and 2mM EDTA) and fixed overnight using 2% formaldehyde made in PBS (VWR, Sweden). The broad extended panel of antibodies used are listed in Supplementary Table 1. For acquisition by CyTOF, cells were stained with DNA intercalator (0.125 μM Iridium-191/193 or MaxPar® Intercalator-Ir, Fluidigm) in 2% formaldehyde made in PBS for 20 min at room temperature. Cells were washed once with CyFACS buffer, PBS and milliQ water, and twice with Cell acquisition solution (CAS) (Fluidigm). Cells were mixed with 0.1X Norm Beads (EQ^TM^ Four Element Calibration Beads, Fluidigm) filtered through a 35µm nylon mesh and diluted to 1000,000 cells/ml. Cells were acquired using Helios mass cytometer at a rate of 300–500 cells/s using PSI system, CyTOF software version 6.5.358 with noise reduction, a lower convolution threshold of 400, event length limits of 10–150 pushes, a sigma value of 3, and flow rate of 0.030 ml/min.

### Antibodies and reagents

Purified antibodies for mass cytometry were obtained in carrier/protein-free buffer and then coupled to lanthanide metals using the MaxPar antibody conjugation kit (Fluidigm Inc.) as per the manufacturer’s recommendations. Following the protein concentration determination by measurement of absorbance at 280 nm on a nanodrop, the metal-labeled antibodies were diluted in Candor PBS Antibody Stabilization solution (Candor Bioscience, Germany) for long-term storage at 4°C. Antibodies used are listed in Supplementary Table 1.

### Plasma protein profiling

Serum or plasma samples collected from patients with COVID-19 (by spinning blood at 2000g for 10min at 80 C for plasma collection or by collecting serum from those blood samples from which PBMCs were isolated using gradient centrifugation for future use and not intended for this study) were analyzed using a multiplex proximity extension assay (OLINK Bioscience, Uppsala, Sweden). Each kit provides a microtiter plate for measuring 92 protein biomarkers. Two panels, the Olink Immune Response and Inflammation panels. Each well contains 96 pairs of DNA-labeled antibody probes. Samples were incubated in the presence of proximity antibody pairs tagged with DNA reporter molecules. When the antibody pair bounds to their corresponding antigens, the corresponding DNA tails form an amplicon by proximity extension, which can be quantified by high-throughput real-time PCR.

### Detection of anti-SARS-CoV-2 antibody response

Antibodies against SARS-CoV-2 were measured using indirect immunofluorescence assay (IFA) and enzyme-linked immunosorbent assay (ELISA) using SARS-CoV-2 receptor-binding domain (RBD) as the antigen. The IFA was conducted as described (Haveri et al., 2020). The RBD ELISA was done following a recently published protocol (Amanat et al., 2020; Stadlbauer et al., 2020). The RBD antigen was produced by transient transfection of RBD plasmid to Vero E6 cells and the produced protein was purified following established protocol (Stadlbauer et al., 2020). The raw data is available in Supplementary Table 2.

## QUANTIFICATION AND STATISTICAL ANALYSIS

### Mass Cytometry Preprocessing and Gating

All FCS-files unrandomized using the CyTOF software (version 6.0.626) were transferred without any additional preprocessing.

### Multiomics Factor Analysis, MOFA

MOFA (Argelaguet et al., 2018) uses a set of data matrices as input, plasma protein expression and cell abundance datasets were used to build the *MOFAobject* with *MultiAssayExperiment*. The MOFA object was trained in R through the *reticulate* package with 10 factors and a variance threshold of 0.01%. Both omics datasets were processed individually to remove any features resulting in zero or low variance before fitting the model. Convergence of the model was assessed using the change in ELBO (deltaELBO) to verify it fit the convergence threshold which is considered to be between 1 and 10. Multiple models were run under different initializations to validate that factors were consistent across trials for model selection.

### Partition-based graph abstraction of single-cell data

The CyTOF data were first preprocessed with arcsin h and scaled to unit variance and then partitioned into different subpopulations according to our in-house supervised learning algorithm. For each subpopulation, the phenotypic changes over different time points are inferred with a trajectory inference method called PAGA (Wolf et al., 2019). In brief, PCA was first applied to reduce the number of features to 20, and then the clusters were detected with Leiden method (Traag et al., 2019). Afterwards, the Leiden output was used by PAGA to infer a trajectory map of clusters. Finally, the PAGA graph was taken as the initial position by ForceAtlas2 (FA)(Jacomy et al., 2014) for the single-cell level visualization.

### Mixed effects modeling

A partially Bayesian method was applied with *blme* package on both datasets (plasma protein expression and cell abundance) to produce maximum a posteriori (MAP) estimates (Chung et al., 2013). This provided the ability to nest the variables, and account for days from admission as well as RBD levels.

## DATA AND SOFTWARE AVAILABILITY

### Raw Mass cytometry data is available for download

https://ki.box.com/s/sby0jesyu23a65cbgv51vpbzqjdmipr1 Olink protein data is presented in Suppl. Table 3.

---

**Suppl. Table 1.** Antibodies used for Mass cytometry.

| Metal tag | Marker | Clone | Vendor |
| --- | --- | --- | --- |
| 89Y | CD45 | HI30 | Fluidigm |
| 110Cd | CD33 | WM53 | Biolegend |
| 111Cd | CD26 | BA5b | Biolegend |
| 112Cd | CD11c | Bu15 | Biolegend |
| 113Cd | IgD | IA6-2 | Biolegend |
| 114Cd | HLA-DR | L243 | BioLegend |
| 115In | CD57 | HCD57 | Biolegend |
| 141Pr | CD49d | 9F10 | Fluidigm |
| 142Nd | CD43 | 84-3C1 | eBiosciences |
| 143Nd | CD3e | UCHT1 | Biolegend |
| 144Nd | CD45RB | MEM-55 | Biolegend |
| 145Nd | CD81 | 5A6 | Biolegend |
| 146Nd | CD52 | HI186 | Biolegend |
| 147Sm | CD1c | L161 | Biolegend |
| 148Nd | CD55 | JS11 | Biolegend |
| 149Sm | CD25 | 2A3 | Fluidigm |
| 150Nd | CD64 | 10.1 | Biolegend |
| 151Eu | CD123 | 6H6 | BioLegend |
| 152Sm | TCRgd | 5A6.E9 | Fischer Scientific |
| 153Eu | Siglec-8 | 837535 | R&D Systems |
| 154Sm | CD95 | DX2 | BioLegend |
| 155Gd | CX3CR1 | 8E10.D9 | BioLegend |
| 156Gd | CD20 | 2H7 | BioLegend |
| 157Gd | CD9 | SN4 C3-3A2 | eBiosciences |
| 158Gd | CD34 | 581 | Biolegend |
| 159Tb | CD22 | HIB22 | Biolegend |
| 160Gd | CD14 | M5E2 | Biolegend |
| 161Dy | CD161 | HP-3G10 | Biolegend |
| 162Dy | CD29 | TS2/16 | Biolegend |
| 163Dy | 4-1BB | 4B4-1 | Biolegend |
| 164Dy | CD62L | DREG-56 | Biolegend |
| 165Ho | CD127 | A019D5 | Fluidigm |
| 166Er | CD24 | ML5 | Biolegend |
| 167Er | CD27 | L128 | Biolegend |
| 168Er | CD141 | M80 | Biolegend |
| 169Tm | CD45RA | HI100 | Fluidigm |
| 170Er | CD38 | HIT2 | Biolegend |
| 171Yb | CD85j | GHI/75 | Biolegend |
| 172Yb | CD147 | HIM6 | BioLegend |
| 173Yb | CD56 | NCAM16.2 | BD Pharmingen |
| 174Yb | CD99 | HCD99 | Biolegend |
| 175Lu | CD28 | CD28.2 | Biolegend |
| 176Yb | CD39 | A1 | Biolegend |
| 191Ir | DNA Ir | Cell-ID DNA Intercalator | Fluidigm |
| 193Ir | DNA Ir | Cell-ID DNA Intercalator | Fluidigm |
| 194Pt | CD8a | SK1 | BD Pharmingen |
| 195Pt | CD5 | UCHT2 | Biolegend |
| 196Pt | CD7 | CD7-6B7 | Biolegend |
| 198Pt | CD4 | RPA-T4 | Biolegend |
| 209Bi | CD16 | 3G8 | Fluidigm |

**Suppl. Table 2.** IgG data.

| Sample ID | Subject ID | IFA (inf. cells) | old-RBD | VE6-RBD |
| --- | --- | --- | --- | --- |
| CoV-24, serum, t1 | COV-24 | 320 | 0.138 | 0.064 |
| CoV-24, heparin, t2 | COV-24 | 2570 | 0.902 | 0.2025 |
| CoV-25, serum, t1 | COV-25 | 320 | 0.219 | 0.0795 |
| CoV-25, EDTA, t2 | COV-25 | 1280 | 0.533 | 0.118 |
| CoV-26, serum, t1 | COV-26 | 2560 | 0.458 | 0.1015 |
| CoV-26, heparin, t2 | COV-26 | 2570 | 0.866 | 0.163 |
| CoV-27, serum, t1 | COV-27 | 640 | 0.666 | 0.1415 |
| CoV-27, heparin, t2 | COV-27 | 1280 | 1.139 | 0.218 |
| CoV-28, serum, t1 | COV-28 | 640 | 0.385 | 0.1 |
| CoV-29, serum, t1 | COV-29 | 160 | 0.21 | 0.085 |
| CoV-29, heparin, t2 | COV-29 | 1280 | 0.923 | 0.1865 |
| CoV-30, serum, t1 | COV-30 | 1280 | 0.928 | 0.1895 |
| CoV-30, serum, t2 | COV-30 | 2570 | 1.063 | 0.215 |
| CoV-31, serum, t1 | COV-31 | 320 | 0.654 | 0.135 |
| CoV-31, serum, t2 | COV-31 | 640 | 0.926 | 0.185 |
| CoV-32, serum, t1 | COV-32 | 80 | 0.084 | 0.0645 |
| CoV-32, heparin, t2 | COV-32 | 1280 | 0.45 | 0.1035 |
| CoV-33, serum, t1 | COV-33 | 80 | 0.17 | 0.0685 |
| CoV-33, heparin, t2 | COV-33 | 640 | 0.559 | 0.112 |
| CoV-34, EDTA, t1 | COV-34 | 10 | 0.077 | 0.0625 |
| CoV-34, EDTA, t2 | COV-34 | 2560 | 1.138 | 0.267 |
| CoV-34, EDTA, t3 | COV-34 | 2560 | 1.035 | 0.237 |
| CoV-35, serum, t1 | COV-35 | 10 | 0.073 | 0.06 |
| CoV-35, heparin, t2 | COV-35 | 2560 | 0.672 | 0.141 |
| CoV-36, serum, t1 | COV-36 | 10 | 0.061 | 0.057 |
| CoV-36, serum, t2 | COV-36 | 640 | 0.485 | 0.117 |
| CoV-37, serum, t1 | COV-37 | 10 | 0.075 | 0.0615 |
| CoV-37, heparin, t2 | COV-37 | 80 | 0.086 | 0.063 |
| CoV-39, serum, t1 | COV-39 | 640 | 0.277 | 0.081 |
| CoV-39, heparin, t2 | COV-39 | 640 | 0.61 | 0.123 |
| CoV-40, serum, t1 | COV-40 | 1280 | 0.084 | 0.065 |
| CoV-40, heparin, t2 | COV-40 | 2570 | 0.909 | 0.2445 |
| CoV-40, EDTA, t3 | COV-40 | 2560 | 0.869 | 0.216 |
| CoV-41, serum, t1 | COV-41 | 320 | 0.081 | 0.0615 |
| CoV-41, heparin, t2 | COV-41 | 640 | 0.308 | 0.0985 |

**Suppl. Table 3. Raw Olink data**

## REFERENCES

Ackermann, M., Verleden, S.E., Kuehnel, M., Haverich, A., Welte, T., Laenger, F., Vanstapel, A., Werlein, C., Stark, H., Tzankov, A., et al. (2020). Pulmonary Vascular Endothelialitis, Thrombosis, and Angiogenesis in Covid-19. New Engl J Medicine.

Amanat, F., Stadlbauer, D., Strohmeier, S., Nguyen, T.H.O., Chromikova, V., McMahon, M., Jiang, K., Arunkumar, G.A., Jurczyszak, D., Polanco, J., et al. (2020). A serological assay to detect SARS-CoV-2 seroconversion in humans. Nat Med 1–4.

Argelaguet, R., Velten, B., Arnol, D., Dietrich, S., Zenz, T., Marioni, J.C., Buettner, F., Huber, W., and Stegle, O. (2018). Multi-Omics Factor Analysis—a framework for unsupervised integration of multi omics data sets. Mol Syst Biol 14, e8124.

Blanco-Melo, D., Nilsson-Payant, B.E., Liu, W.-C., Uhl, S., Hoagland, D., Møller, R., Jordan, T.X., Oishi, K., Panis, M., Sachs, D., et al. (n.d.). Imbalanced host response to SARS-CoV-2 drives development of COVID-19. Cell.

Brodin, P. (2018). The biology of the cell – insights from mass cytometry. Febs J.

Brodin, P. (2020). Why is COVID-19 so mild in children? Acta Paediatr.

Brodin, P., and Davis, M.M. (2016). Human immune system variation. Nat Rev Immunol 17, 21–29.

Brodin, P., Jojic, V., Gao, T., Bhattacharya, S., Angel, C.J.L., Furman, D., Shen-Orr, S., Dekker, C.L., Swan, G.E., Butte, A.J., et al. (2015). Variation in the Human Immune System Is Largely Driven by Non-Heritable Influences. Cell 160, 37–47.

Brodin, P., Duffy, D., and Quintana-Murci, L. (2019). A Call for Blood—In Human Immunology. Immunity 50, 1335–1336.

Casanova, J.-L., Su, H.C., and Effort, C.H.G. (2020). A global effort to define the human genetics of protective immunity to SARS-CoV-2 infection. Cell.

Chung, Y., Rabe-Hesketh, S., Dorie, V., Gelman, A., and Liu, J. (2013). A Nondegenerate Penalized Likelihood Estimator for Variance Parameters in Multilevel Models. Psychometrika 78, 685–709.

Denzel, A., Maus, U.A., Gomez, M.R., Moll, C., Niedermeier, M., Winter, C., Maus, R., Hollingshead, S., Briles, D.E., Kunz-Schughart, L.A., et al. (2008). Basophils enhance immunological memory responses. Nat Immunol 9, 733–742.

Flores-Torres, A.S., Salinas-Carmona, M.C., Salinas, E., and Rosas-Taraco, A.G. (2019). Eosinophils and Respiratory Viruses. Viral Immunol 32, 198–207.

Grifoni, A., Weiskopf, D., Ramirez, S.I., Mateus, J., Dan, J.M., Moderbacher, C.R., Rawlings, S.A., Sutherland, A., Premkumar, L., Jadi, R.S., et al. (2020). Targets of T cell responses to SARS-CoV-2 coronavirus in humans with COVID-19 disease and unexposed individuals. Cell.

Grishkan, I.V., Ntranos, A., Calabresi, P.A., and Gocke, A.R. (2013). Helper T cells down-regulate CD4 expression upon chronic stimulation giving rise to double-negative T cells. Cell Immunol 284, 68–74.

Haveri, A., Smura, T., Kuivanen, S., Österlund, P., Hepojoki, J., Ikonen, N., Pitkäpaasi, M., Blomqvist, S., Rönkkö, E., Kantele, A., et al. (2020). Serological and molecular findings during SARS-CoV-2 infection: the first case study in Finland, January to February 2020. Eurosurveillance 25, 2000266.

Huang, C., Wang, Y., Li, X., Ren, L., Zhao, J., Hu, Y., Zhang, L., Fan, G., Xu, J., Gu, X., et al. (2020). Clinical features of patients infected with 2019 novel coronavirus in Wuhan, China. Lancet 395, 497–506.

Iwasaki, A., and Yang, Y. (2020). The potential danger of suboptimal antibody responses in COVID-19. Nat Rev Immunol 1–3.

Jacomy, M., Venturini, T., Heymann, S., and Bastian, M. (2014). ForceAtlas2, a continuous graph layout algorithm for handy network visualization designed for the Gephi software. Plos One 9, e98679.

Jin, J.-M., Bai, P., He, W., Wu, F., Liu, X.-F., Han, D.-M., Liu, S., and Yang, J.-K. (2020). Gender Differences in Patients With COVID-19: Focus on Severity and Mortality. Frontiers Public Heal 8, 152.

Kawakami, T. (2008). Basophils now enhance memory. Nat Immunol 9, 720–721.

Lagunas-Rangel, F.A. (2020). Neutrophil-to-lymphocyte ratio and lymphocyte-to-C-reactive protein ratio in patients with severe coronavirus disease 2019 (COVID-19): A meta-analysis. J Med Virol.

Lakshmikanth, T., Muhammad, S.A., Olin, A., Chen, Y., Mikes, J., Fagerberg, L., Gummesson, A., Bergström, G., Uhlen, M., and Brodin, P. (2020). Human immune system variation during one year. Biorxiv 2020.01.22.915025.

Lazear, H.M., Schoggins, J.W., and Diamond, M.S. (2019). Shared and Distinct Functions of Type I and Type III Interferons. Immunity 50, 907–923.

Lu, R., Zhao, X., Li, J., Niu, P., Yang, B., Wu, H., Wang, W., Song, H., Huang, B., Zhu, N., et al. (2020). Genomic characterisation and epidemiology of 2019 novel coronavirus: implications for virus origins and receptor binding. Lancet 395, 565–574.

Lundberg, M., Eriksson, A., Tran, B., Assarsson, E., and Fredriksson, S. (2011). Homogeneous antibody-based proximity extension assays provide sensitive and specific detection of low-abundant proteins in human blood. Nucleic Acids Res 39, e102–e102.

Mathew, D., Giles, J.R., Baxter, A.E., Greenplate, A.R., Wu, J.E., Alanio, C., Oldridge, D.A., Kuri-Cervantes, L., Pampena, M.B., D’Andrea, K., et al. (2020). Deep immune profiling of COVID-19 patients reveals patient heterogeneity and distinct immunotypes with implications for therapeutic interventions. Biorxiv 2020.05.20.106401.

Medzhitov, R., and Janeway, C.A. (2002). Decoding the Patterns of Self and Nonself by the Innate Immune System. Science 296, 298–300.

Mesnil, C., Raulier, S., Paulissen, G., Xiao, X., Birrell, M.A., Pirottin, D., Janss, T., Starkl, P., Ramery, E., Henket, M., et al. (2016). Lung-resident eosinophils represent a distinct regulatory eosinophil subset. J Clin Investigation 126, 3279–3295.

Mikes, J., Olin, A., Lakshmikanth, T., Chen, Y., and Brodin, P. (2019). Mass Cytometry, Methods and Protocols. Methods Mol Biology Clifton N J 1989, 111–123.

Momose, T., Okubo, Y., Horie, S., Takashi, S., Tsukadaira, A., Suzuki, J., Isobe, M., and Sekiguchi, M. (1999). Interferon-γ Increases CD62L Expression on Human Eosinophils. Int Arch Allergy Imm 120, 30–33.

Olin, A., Henckel, E., Chen, Y., Lakshmikanth, T., Pou, C., Mikes, J., Gustafsson, A., Bernhardsson, A.K., Zhang, C., Bohlin, K., et al. (2018). Stereotypic Immune System Development in Newborn Children. Cell 174, 1277–1292.e14.

Robbiani, D.F., Gaebler, C., Muecksch, F., Lorenzi, J.C.C., Wang, Z., Cho, A., Agudelo, M., Barnes, C.O., Gazumyan, A., Finkin, S., et al. (2020). Convergent Antibody Responses to SARS-CoV-2 Infection in Convalescent Individuals. Biorxiv 2020.05.13.092619.

Stadlbauer, D., Amanat, F., Chromikova, V., Jiang, K., Strohmeier, S., Arunkumar, G.A., Tan, J., Bhavsar, D., Capuano, C., Kirkpatrick, E., et al. (2020). SARS-CoV-2 Seroconversion in Humans: A Detailed Protocol for a Serological Assay, Antigen Production, and Test Setup. Curr Protoc Microbiol 57, e100.

Sun, B., Feng, Y., Mo, X., Zheng, P., Wang, Q., Li, P., Peng, P., Liu, X., Chen, Z., Huang, H., et al. (2020). Kinetics of SARS-CoV-2 specific IgM and IgG responses in COVID-19 patients. Emerg Microbes Infec 9, 940–948.

Taniguchi, H., Katoh, S., Kadota, J., Matsubara, Y., Fukushima, K., Mukae, H., Matsukura, S., and Kohno, S. (2000). Interleukin 5 and granulocyte-macrophage colony-stimulating factor levels in bronchoalveolar lavage fluid in interstitial lung disease. Eur Respir J 16, 959–964.

Tetro, J.A. (2020). Is COVID-19 receiving ADE from other coronaviruses? Microbes Infect 22, 72–73.

Traag, V.A., Waltman, L., and Eck, N.J. van (2019). From Louvain to Leiden: guaranteeing well-connected communities. Sci Rep-Uk 9, 5233.

Uhlen, M., Karlsson, M.J., Zhong, W., Tebani, A., Pou, C., Mikes, J., Lakshmikanth, T., Forsström, B., Edfors, F., Odeberg, J., et al. (2019). A genome-wide transcriptomic analysis of protein-coding genes in human blood cells. Sci New York N Y 366.

Vabret, N., Britton, G.J., Gruber, C., Hegde, S., Kim, J., Kuksin, M., Levantovsky, R., Malle, L., Moreira, A., Park, M.D., et al. (2020). Immunology of COVID-19: current state of the science. Immunity.

Vardhana, S.A., and Wolchok, J.D. (2020). The many faces of the anti-COVID immune response. J Exp Medicine 217.

Wolf, F.A., Hamey, F.K., Plass, M., Solana, J., Dahlin, J.S., Göttgens, B., Rajewsky, N., Simon, L., and Theis, F.J. (2019). PAGA: graph abstraction reconciles clustering with trajectory inference through a topology preserving map of single cells. Genome Biol 20, 59.

Xiang, J., McLinden, J.H., Rydze, R.A., Chang, Q., Kaufman, T.M., Klinzman, D., and Stapleton, J.T. (2009). Viruses within the Flaviviridae decrease CD4 expression and inhibit HIV replication in human CD4+ cells. J Immunol Baltim Md 1950 183, 7860–7869.

Zhou, Y., Fu, B., Zheng, X., Wang, D., Zhao, C., qi, Y., Sun, R., Tian, Z., Xu, X., and Wei, H. (2020). Aberrant pathogenic GM-CSF+ T cells and inflammatory CD14+CD16+ monocytes in severe pulmonary syndrome patients of a new coronavirus. Biorxiv 2020.02.12.945576.

